# Young adults with recurrent low back pain demonstrate altered trunk coordination during gait independent of pain status and attentional demands

**DOI:** 10.1101/2020.09.28.20203208

**Authors:** Hai-Jung Steffi Shih, Carolee J. Winstein, Kornelia Kulig

## Abstract

Pain influences both attention and motor behavior. We used a dual-task interference paradigm to investigate 1) alterations in attentional performance, 2) the ability to switch task prioritization, and 3) the effect of attentional demand on trunk coordination during narrow-based walking in and out of a painful episode in individuals with recurrent low back pain (LBP). We tested twenty young adults with LBP both in and out of a painful episode and compared them to twenty matched back-healthy individuals. Participants simultaneously performed a narrow step width matching task and an arithmetic task, with and without instructions to prioritize either task. A motion capture system was used to record kinematic data, and frontal plane trunk coordination was analyzed using vector coding on the thorax and pelvis angles. Single task performance, dual-task effect, dual-task performance variability, task prioritization switch, and trunk coordination were analyzed using paired t-tests or repeated measures two-way ANOVAs. Results indicated that active pain has a detrimental effect on attentional processes, indicated by poorer single task performance and increased dual-task performance variability for individuals with recurrent LBP. Individuals with LBP, regardless of pain status, were able to switch task prioritization to a similar degree as their back-healthy counterparts. Compared to the control group, individuals with recurrent LBP exhibited a less in-phase, more pelvis-dominated trunk coordination during narrow-based walking, independent of pain status and regardless of attentional manipulations. Thus, altered trunk coordination in persons with LBP appears to be habitual, automatic, and persists beyond symptom duration.

## Introduction

Pain is a potent stimulus that captures attention, occupies executive resources, and impacts dual-task performance [1–3]. Pain associated with movement may elicit a greater degree of executive control in typically less attention-demanding movements (e.g. walking) to minimize additional discomfort [4]. Further, the ability to prioritize tasks under dual-task conditions may be compromised in the presence of movement-related pain. The simultaneous implementation of a postural task and a cognitive task requires appropriate allocation of attentional resources to foster safe and successful execution. Previous literature has demonstrated that as postural challenge increases, priority shifts towards the postural task over the cognitive task [5]. This effect may be even more profound in individuals with decreased postural reserve [6], or with increased risk associated with the motor task such as movement-related pain.

Low back pain (LBP) is usually movement-provoked; as such LBP serves as a reasonable model to investigate the influence of pain on attention and task-prioritization during dual-task walking. Previous studies suggest altered motor behavior under dual-task conditions in individuals with LBP. There is emerging evidence that spatiotemporal gait variability [7,8], trunk movement and coordination variability [9,10], and postural control [3,11,12] are differentially affected in individuals with LBP compared to controls when under dual-task conditions. Previously, trunk coordination in the transverse and frontal planes was shown to be more in-phase and coupled between thorax and pelvis during single-task walking in individuals with LBP, perhaps to minimize movement between spinal segments and reduce irritation to the surrounding tissues [13–15]. However, studies that used a dual-task paradigm were inconsistent in their findings — two studies reported no difference between groups [8,9] and one study reported an increase of in-phase coordination in the LBP group under dual-task interference [16].

Dual-task designs used in previous research often lack a comprehensive evaluation of both the motor and cognitive task performance, a critical limitation that overlooks the well-known trade-off between tasks due to cognitive-motor interference [17,18]. Cognitive-motor intereference often causes both motor and cognitive task performance to change from a single-task condition, however the magnitude and direction of the change could be impacted by individual factors such as age, task competency/familiarity, cognitive ability, and task prioritization/preference. Therefore, without an explicit manipulation of task priority, the precise dual-task effects are uninterpretable — is the performance decrement due to pain (group comparison), interference (dual-task vs single task), and/or task priority (implicit vs explicit)? Furthermore, individuals with LBP may naturally prioritize the motor task given the perceived risk of inducing pain if the movements are not executed a certain way. By introducing explicit prioritization instructions, we will gain insight into whether individuals with LBP can switch task prioritization between the cognitive and motor tasks to the same degree as back-healthy individuals.

Neuroplasticity and learning may drive persistent changes in dual-task performance when pain becomes long-term. There is evidence of alterations in functional connectivity and white matter anatomy in the dorsal-lateral prefrontal cortex, part of the cognitive/attentional network, in individuals with chronic LBP [19,20]. Some dual-task studies tested participants with LBP during a painful episode [7,9–11], while others tested them during symptom remission [8,12], but their results were too dispersed to draw conclusions. Whether changes to dual-task performance is solely due to active pain, or if there is a persistent impact on attentional resources beyond symptom duration is an important question that we aim to answer with the current study. Motivated by previous reports of limited recovery of brain morphology after pain subsides [21] and residual motor behavior alterations in asymptomatic individuals with a history of pain [8,16,22], we expected the impact of an active pain episode on dual-task performance and trunk coordination to persist beyond symptom duration in those with recurrent LBP.

We designed this study using a dual-task interference paradigm to investigate 1) attention (assessed by task performance), 2) task prioritization, and 3) the effect of attentional demand on trunk coordination during gait in and out of a painful episode in a cohort of individuals with recurrent LBP and to compare them to back-healthy individuals. By doing so we addressed two unanswered questions – first, can individuals with LBP switch task prioritization between cognitive and motor tasks when instructed, and second, do alterations in attentional processes persist both in and out of LBP episodes? We hypothesized that attention (measured by dual-task performance and performance variability) and the ability to voluntarily switch task priorities during a challenging walking task (i.e., narrow-based walking) will be diminished in individuals with recurrent LBP, regardless of pain status, compared to their back-healthy counterparts. Further, we tested whether individuals with recurrent LBP, regardless of pain status, exhibit more in-phase trunk coordination than controls, across single and dual-task conditions.

## Methods

### Participants

Based on pilot data, our sample size calculation suggested that at least 16 participants in each group would be sufficient to reach 80% statistical power to detect differences in 1) step width dual-task performance, 2) change in step width performance between prioritization instructions, and 3) trunk coordination patterns. Twenty young adults with recurrent LBP and twenty age, sex, BMI, and activity-matched back-healthy individuals participated. Participants with recurrent LBP were included if they are between 18 to 45 years old, have activity-limiting back pain for more than 6 months, but had less than half of the days in pain. Back-healthy participants were included if they had no back pain in the previous year. Participants were excluded if they had a history of leg pain below the knee accompanying back pain, have chronic or recurrent pain in other body regions, a history of low back, lower extremity, or cervical surgery, any known spinal fracture or pathology, a history of diabetes mellitus, active cancer, current pregnancy, or any condition that is known to affect balance or locomotion. To eliminate potential effects of stimulants, they were also excluded if they consume alcohol for more than 10 drinks per week, caffeinated drink for more than 4 cups per day, or tobacco for more than 15 cigarettes per day.

Participants with recurrent LBP were tested twice, first when their pain persisted for more than 24 hours at the level of ≥2/10 on the numeric rating scale [23], and then again when their pain is <1/10 on the numeric rating scale for more than 24 hours. Given the high test-retest reliability in the pilot control group, back-healthy individuals were only tested once. Participants provided written informed consent that was approved by the University of Southern California institutional review board.

### Instrumentation

Participants were instrumented with a typical lower extremity marker-set, with additional markers placed on the pelvis and thorax including anterior superior iliac spines, iliac crest, and S1, bilateral acromion, sternal notch, and T1. Kinematic data were recorded by a 11-camera Qualysis motion capture system (Qualisys Inc., Gothenburg, Sweden) at 125 Hz. A portable treadmill (PRO-FORM 505 CST, ICON Health & Fitness, Inc., Logan, UT, USA) was used for the walking trials.

### Experimental procedures

Participants were screened for scoliosis and radiculopathy, and completed a medical history form, the International Physical Activity Questionnaire (IPAQ) [24,25], and the Montreal Cognitive Assessment (MoCA) [26]. Individuals with recurrent LBP also completed the Oswestry Disability Index (ODI) [27].

For all treadmill walking trials, the speed was set at 1.25 m/s. Participants were given 3 minutes to familiarize with the treadmill, after which their preferred step width was determined by a 30 second treadmill walking trial. Participants were then introduced to the step width real-time visual feedback projected on the wall in front of the treadmill (Fig 1A). Step width was calculated using marker data that was streamed real-time into MATLAB (MathWorks, Inc., Natick, MA, USA) by finding the medial-lateral distance between the averaged heel and 2^nd^ toe marker position every foot flat. Participants were instructed to match their current step width to the target width indicated on the visual feedback as accurately and consistently as possible. The target step width was prescribed as 0.33 times their preferred step width. They were also introduced to the arithmetic task, which required subtracting 7s continuously from a random three-digit number as fast and as accurately as possible. Participants completed five 30-second step-width matching practice trials, three 30-second serial subtraction practice trials while seated, and one dual-task familiarization trial in which they perform the two tasks simultaneously. They then performed under 5 different conditions, 3 trials each: arithmetic single task (seated), step width single task, dual-task (without prioritization instructions), step width prioritization (SW-pri), and arithmetic prioritization (Ari-pri). During the prioritization trials, participants were instructed to do their best on either the step width task or arithmetic task while performing them concurrently. The single and dual-task conditions were randomized and performed first, before the two prioritization conditions were randomized and administered to avoid contamination of the prioritization condition to the no-priority-instructed dual-task condition.

**Figure 1.**
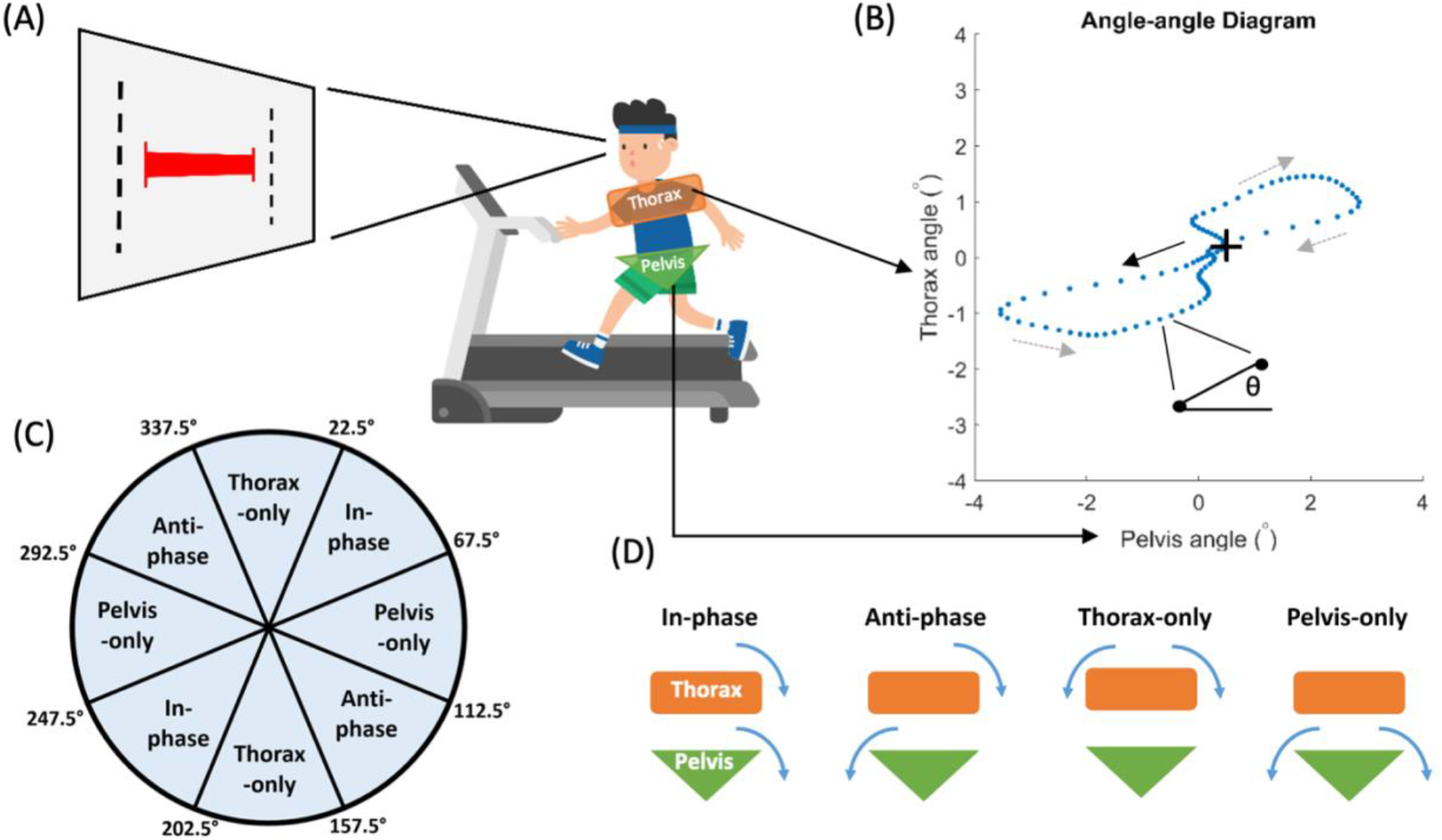
(A) Experimental setup of treadmill walking while matching a narrow step width. A visual feedback was projected on a wall in front of the treadmill, with a red horizontal bar representing participant’s actual step width and black vertical lines indicating the target width. (B) Pelvis-thorax angle-angle diagram of a gait cycle in one representative participant. The “+” denotes right heel initial contact and the arrows indicate direction of progression of movement. Coupling angleθ was defined as the vector angle of two consecutive points in time relative to the right horizontal. (C) Cutoffs for binning of the coupling angles into four coordination patterns. (D) Illustrations of the four trunk coordination patterns. In-phase indicates that both segments are rotating towards the same direction at similar rate; anti-phase indicates that the segments are rotating to the opposite direction at similar rate; thorax-only and pelvis-only indicate that the thorax or pelvis segment is rotating significantly faster than the other segment, or the other segment is hardly rotating.

### Data Analyses

Task performance of the step width task was assessed using root mean square error of participant’s actual step width relative to the target width. Performance of the arithmetic task was calculated as the rate of correct response (the number of correct response where they correctly subtracted 7 from their last answer divided by trial length which was 30 seconds). Since there was a positive relationship between single task performance and the magnitude of change from single to dual-task performance, we calculated the dual-task effect 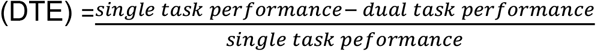 for both tasks to control for single task differences while assessing the impact of dual-task on participant’s performance. There was no relationship between single task performance and the change of task performance between prioritization instructions, therefore we assessed the effect of task prioritization instructions using the raw difference in task performance between step width- and arithmetic-prioritization conditions (ΔSW performance and ΔArithmetic performance). Dual-task performance variability was calculated as coefficient of variation (CV) of task performance under the different dual-task conditions from each individual.

Kinematic data were low-pass filtered at 10 Hz with a dual-pass 4^th^ order Butterworth filter. The frontal plane trunk kinematic coordination was chosen because of its sensitivity in response to narrow step width [28], and was calculated using vector coding analysis described by Needham et al [29]. An angle-angle diagram was first constructed, then coupling angle was determined as the angle of the vector between two adjacent data points in time relative to the right horizontal (Fig 1B). Coordination patterns were categorized as in-phase, anti-phase, thorax-only, and pelvic-only defined by coupling angles falling within each range indicated in Fig 1C (Fig 1C&D).

### Statistical Analysis

Descriptive statistics were performed, and paired t-tests were used to compare participant demographics. To examine our first aim, whether attention was affected by pain or a history of pain, single task performance, dual-task effects, and dual-task performance variability were analyzed. For the second aim where we assess task prioritization, ΔSW performance and ΔArithmetic performance were analyzed. For the third aim focusing on the effect of attentional demand on trunk coordination during gait, the four patterns of trunk coordination were analyzed. These dependent variables were compared between pain status within the LBP group and between the LBP and control groups using the following steps. First, to examine if pain status affects behavior, the dependent variables are compared within the LBP group when they were in active pain (LBP-A) and in remission (LBP-R). Paired t-tests will be used on single task performance, dual-task effect, and changes in task performance under different prioritization instructions. Two-way repeated-measures analysis of variance (ANOVA) were used for dual-task performance variability and trunk coordination, with a pain status by dual-task condition interaction term in the model. If the pain status effect on behavior is not reliable, data for LBP-A and LBP-R will be pooled and compared to the back-healthy controls (CTRL). On the other hand, if there is a significant pain status effect, then LBP-A and LBP-R will be compared separately with CTRL. Of note, if the pain status effect is found not to be reliable, this would provide support for our persistence hypothesis within the LBP group. A main effect of group would allow us to confirm our hypotheses regarding alterations in motor behavior in those with recurrent LBP. A main effect of condition would indicate the effect of dual-task manipulation on behavior across groups, whereas a significant interaction effect would indicate that there is a differential effect of dual-task manipulation on those behaviors in the LBP-A, LBP-R, or pooled LBP vs. CTRL. For all significant main or interaction effects from the ANOVA, a Tukey’s test for post-hoc pairwise comparison will be used to determine the locus of the effect. The α level was set at 0.05. Statistical analyses were done in R [30].

### Reliability and effects of previous task exposure

Six back-healthy participants were re-tested at least one week apart to determine test-retest reliability and to assess potential effects of previous task exposure. For test-retest reliability, intraclass correlation coefficient for one-way random effects, absolute agreement, and multiple measurements (ICC (1,k)) was calculated for step width and arithmetic DTE and for trunk coordination [31]. To assess whether difference between pain status in individuals with recurrent LBP were merely due to previous task exposure, we planned a post-hoc analyses on any variable that was different in and out of pain by performing (1) paired t-tests comparing the test-retest sessions in the controls and (2) Pearson’s correlation between the time to re-test and changes in performance between LBP-A and LBP-R testing sessions. If there was an effect of previous exposure, we would expect (1) an improvement in performance, and (2) the closer the two sessions were in time, the greater improvement in performance (i.e. significant performance gain by time to re-test correlation).

## Results

### Participant characteristics

Per design, no significant differences were found for participants’ demographics between LBP and CTRL groups (Table 1). Back pain characteristics in the LBP group showed that there were significant differences in and out of pain for ODI, pain at rest, and pain during gait (Table 2). Participants with LBP returned for testing in remission after 47.9 ± 44.2 (range = 8-172) days, and the last time they recalled having pain was 10 ± 6.9 (range = 1-23) days earlier.

**Table 1.** Participant demographics (mean ± standard deviation (range)). MoCA: Montreal Cognitive Assessment.

|  | **LBP (n=20)** | **Control (n=20)** | **P-value** |
| --- | --- | --- | --- |
| **Sex** | 6 M, 14 F | 6 M, 14 F | -- |
| **Age (years)** | 25.3 ± 5.2 (19-37) | 26.3 ± 3.3 (22-33) | 0.192 |
| **Height (cm)** | 167.0 ± 7.8 (152-183) | 165.5 ± 9.9 (152-188) | 0.534 |
| **Weight (kg)** | 64.2 ± 12.4 (44-97) | 61.4 ± 12.7 (44-85) | 0.244 |
| **BMI (kg/m<sup>2</sup>)** | 23.0 ± 4.0 (16-31) | 22.2 ± 2.8 (18-29) | 0.132 |
| **Activity (MET)** | 3216 ± 2991 (490-12024) | 2590 ± 1450 (853-6564) | 0.393 |
| **MoCA (0-30)** | 28.80 ± 1.11 (27-30) | 28.65 ± 1.67 (27-30) | 0.651 |

**Table 2.** Mean ± standard deviation (range) for low back pain characteristics. ODI: Oswestry Disability Index; VAS: Visual Analog Scale. Significant p-values are in bold.

|  | **Active Pain** | **In remission** | **P-value** |
| --- | --- | --- | --- |
| **LBP duration (years)** | 5.8 ± 6.2 (0.58-18) | -- | -- |
| **ODI (0-100)** | 17.7 ± 7.9 (8-32) | 4.7 ± 7.9 (0-16) | **<0.001** |
| **Pain at rest VAS (mm) (0-100)** | 40.3 ± 17.5 (21-72) | 1.4 ± 2.4 (0-7) | **<0.001** |
| **Pain during gait VAS (mm) (0-100)** | 36.3 ± 20.6 (0-64) | 1.7 ± 2.2 (0-7) | **<0.001** |

### Attention – assessed by task performance

LBP-A performed worse on the arithmetic single task then LBP-R (p=0.044) and when compared to CTRL (p=0.040) (Fig 2). LBP-A also performed worse on the step width single task then LBP-R(p=0.006), however, their performance both in and out of pain did not differ from the CTRL group (Fig 2).

**Figure 2.**
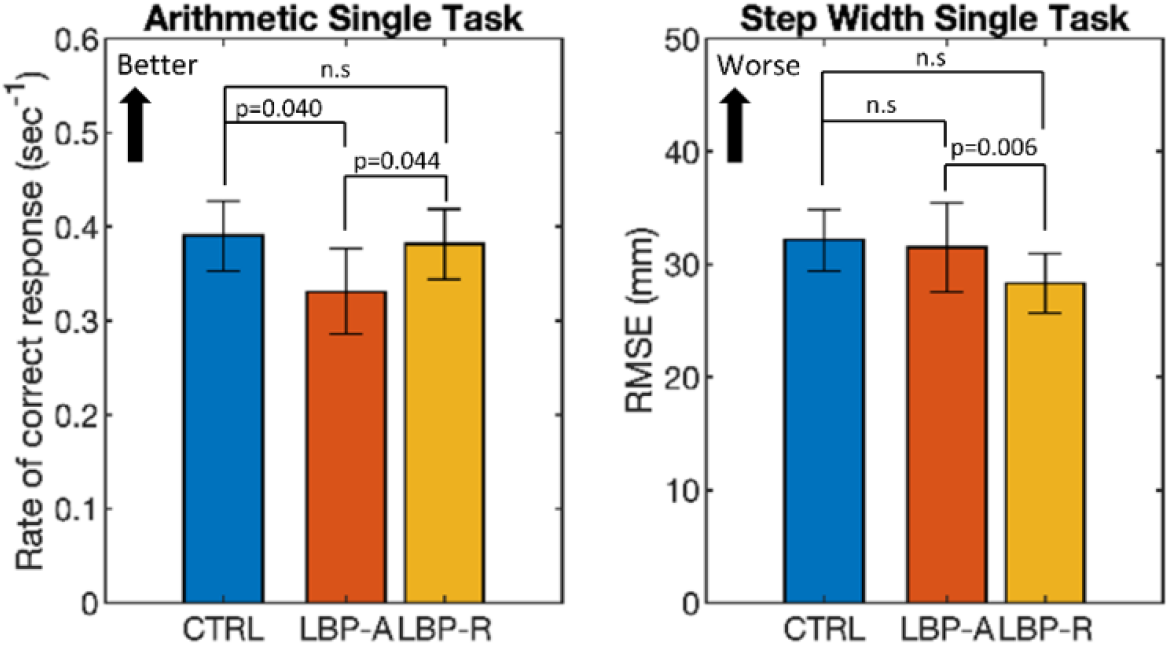
Single task arithmetic and step width performance for the back-healthy controls (CTRL), individuals with recurrent low back pain while in active pain (LBP-A), and the same individuals during symptom remission (LBP-R). Error bars indicate standard error of mean. n.s. = Non-significant.

There were no statistical differences between pain status or groups on arithmetic and step width dual-task effect (Fig 3 A&B). Most individuals had decreased task performance up to 40% for the arithmetic task and 250% for the step width task when dual-tasking. However, there were also a few participants who actually performed better in the dual-task condition.

**Figure 3.**
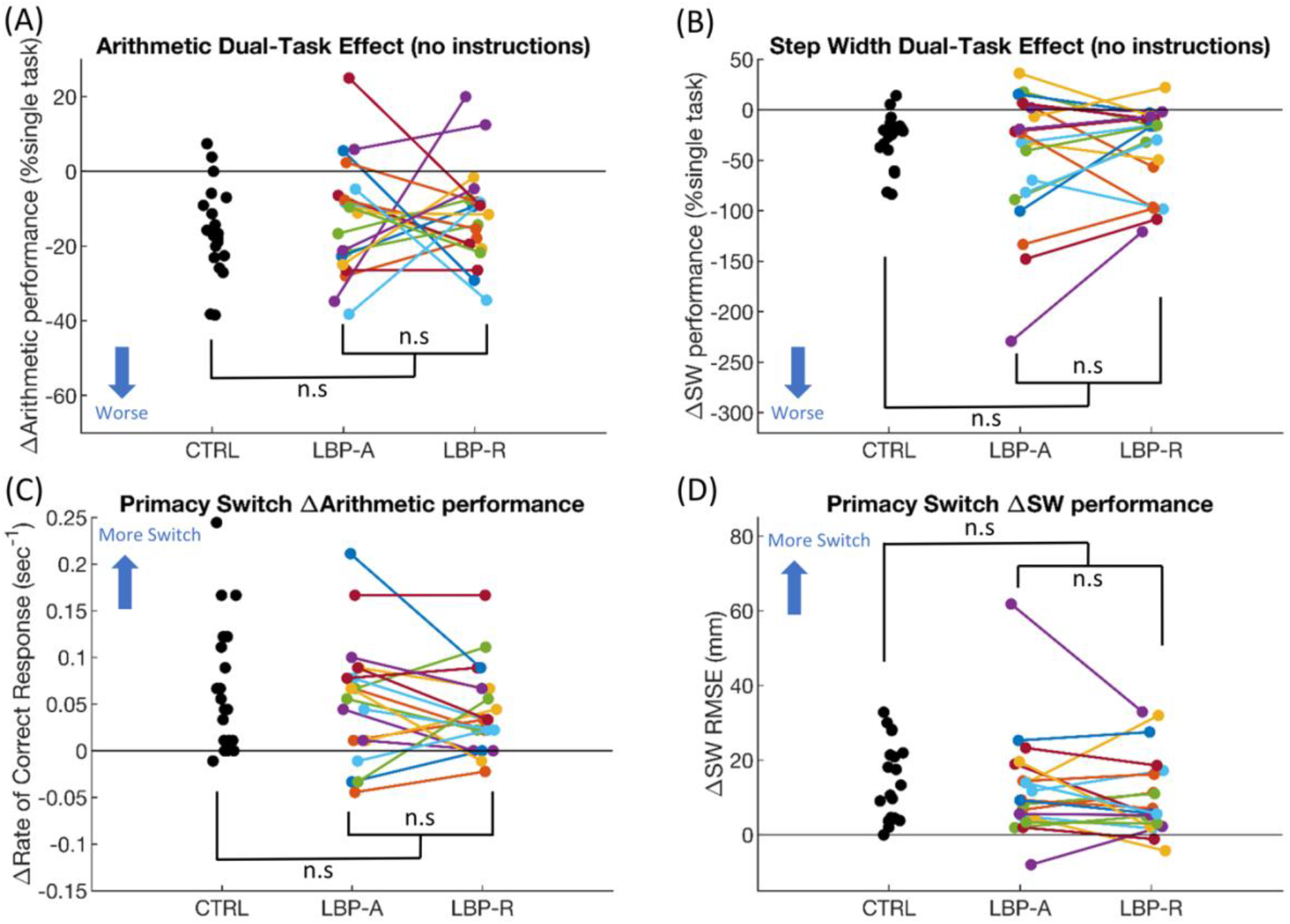
Dual-task performance and primacy switch effects on the control group (CTRL) and the low back pain group in pain (LBP-A) and out of pain (LBP-R). Each dot and each pair of connected dots indicate a single participant, and the reference line at zero would indicate no change in performance. (A) Arithmetic dual-task effect during no instruction condition. (B) Step width dual-task effect during no instruction condition. (C) The change of arithmetic performance from step width-prioritization to arithmetic prioritization conditions. (D) The change of step width performance from step width prioritization to arithmetic prioritization conditions. n.s. = Non-significant.

Results of dual-task performance variability showed that LBP-A had greater variability for both arithmetic and step width tasks than when they were in remission (Fig 4). For arithmetic dual-task performance, the LBP-R had less variability compared to the CTRL. However, for step width dual-task performance there was a trend that the LBP-A had more variability of step width performance compared to the CTRL. There was no main effect of condition, indicating no effect of prioritization instructions on dual-task performance variability. There were also no pain/group x condition interactions, indicating that the effect of pain status and groups were not different across prioritization instructions.

**Figure 4.**
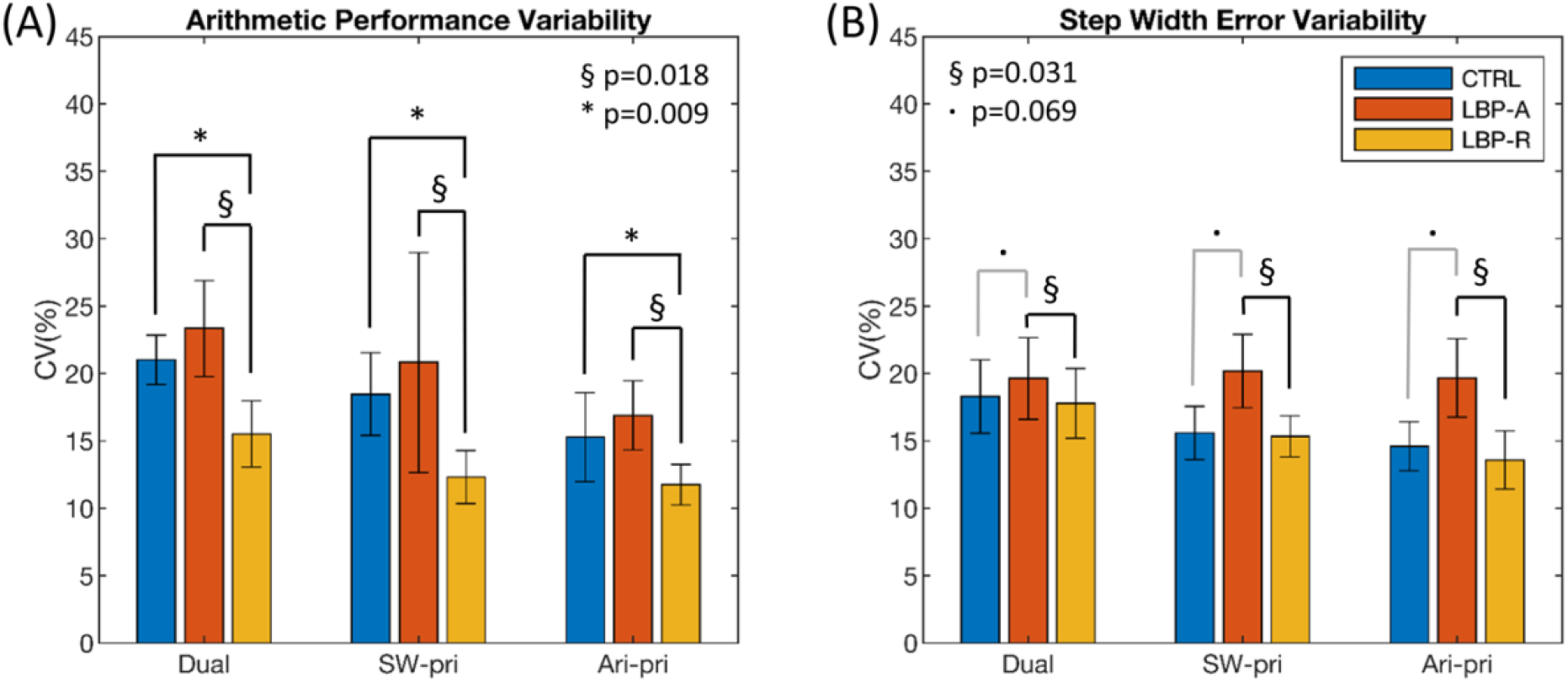
(A) Arithmetic and (B) step width task performance variability during the three dual task conditions shown as coefficient of variation (CV). §: indicates a pain main effect between individuals with recurrent low back pain when they were in (LBP-A) and out of pain (LBP-R). *: indicates a group main effect between back-healthy participants (CTRL) and LBP-R. · : indicates a trend towards significance between CTRL and LBP-A.

### Task prioritization

There were no statistical differences between LBP-A and LBP-R, or pooled LBP and CTRL groups on the effect of prioritization switch on arithmetic and step width task performance (Fig 3 C&D). The majority of the participants in both groups were effectively able to voluntarily switch task priority when instructed. Note the data points lie generally above zero-line in Fig 3C and D, compared to that in Fig 3A and B.

### Trunk coordination

Regardless of pain status, individuals with LBP exhibited trunk coordination patterns that appeared “looser” and more pelvis-dominated during gait compared to controls, and this was not affected by single or dual-task prioritization conditions (Fig 5). There was no difference between LBP-A and LBP-R in any of the four coordination patterns, therefore the coordination results for the LBP group were pooled and compared with the control group. There were main effects of group indicating greater pelvis-only and less in-phase and thorax-only patterns in LBP compared to CTRL. There were also main effects of condition revealing less anti-phase pattern in single task compared to the dual-task conditions with various prioritization instructions, and more pelvis-only pattern in single task compared to the no-priority-instructed dual-task condition. There was no pain/group x condition interaction for any of the comparisons (p>0.05), suggesting that the differences between groups were not influenced by single, dual-task, or different prioritization conditions.

**Figure 5.**
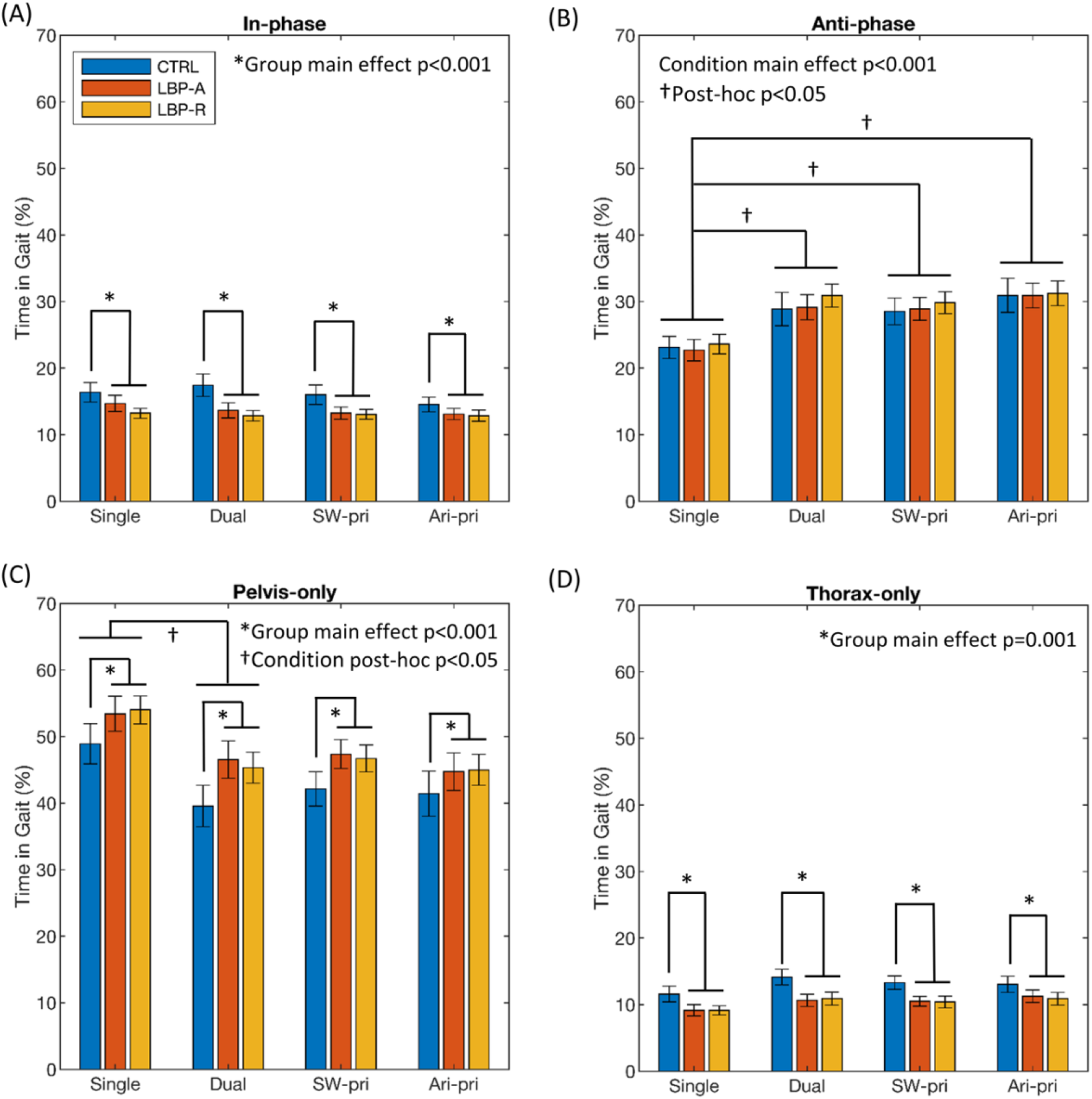
Trunk coordination patterns during single task, dual task (no instruction), step width prioritization (SW-pri) and arithmetic prioritization (Ari-pri) conditions in the control group (CTRL) and the low back pain group in pain (LBP-A) and out of pain (LBP-R). Error bars indicate standard error of mean.

### Reliability and effects of previous task exposure

Test-retest reliability for step width dual-task effect was moderate (ICC=0.668), similar to the ICCs reported for gait speed dual-task effect [32]. Reliability for arithmetic DTE was poor (ICC=0.232), consistent with a previous report of cognitive task ICCs, and may be due to the novelty and the challenge of cognitive tasks [32]. The ICC for the four patterns in trunk coordination ranged between 0.78 and 0.93, representing good to excellent reliability.

Arithmetic and step width single task performance and dual-task performance variability were the only variables that were different between LBP-A and LBP-R. There were no differences between test-retest of the controls in any of those variables (p=0.172-0.953), and there was no relationship between time to re-test and the difference in any of the performance variables in the between LBP-A and LBP-R (Pearson’s r=-0.14-0.23, p=0.280-0.708), indicating no effect of previous exposure to the task (Supplementary Figures 1&2).

## Discussion

We examined dual-task performance, task prioritization, and the effect of attentional demand on trunk coordination during narrow-based walking in young adults with recurrent LBP. For the first time, this study examined the ability of individuals with LBP to switch task prioritization between cognitive and motor tasks when instructed, and the persistence of alterations in attentional processes beyond the painful episode. We found that participants with LBP had altered attention when they were in pain indicated by decreased single task performance and increased dual-task performance variability compared to when they were in symptom remission. Dual-task performance and the ability to switch task prioritization were not different in and out of pain in individuals with recurrent LBP, nor when compared to their back-healthy counterparts, a finding that was inconsistent with our hypothesis. Additionally, individuals with LBP exhibited more pelvis-only coordination and less in-phase and thorax-only patterns than the control group, regardless of whether they were in pain or under single or dual-task conditions. This indicates that the altered trunk coordination patterns in young adults with recurrent LBP were independent of pain status and did not require additional attentional resources, which suggests a habitual, automatic, and persistent movement strategy.

Active pain negatively affected task performance, indicating a detrimental influence on attention. Our results of declined arithmetic and step width single task performance in individuals in an episode of LBP was consistent with previous studies that found decreased cognitive task performance in persons with active LBP [9,11], but no difference in task performance in asymptomatic participants with a history of LBP compared to the control group [8]. An fMRI study had also shown that patients with chronic LBP have decreased activation of the cingulo-frontal-parietal brain network during an attention-demanding task [19]. Additionally, our findings indicate that participants with recurrent LBP were less *consistent* in their dual-task performance when they were in a painful episode than when they were in symptom remission. One viable explanation for these variability results is that active pain may have introduced noise into the sensorimotor system thereby altering the signal to noise ratio and attenuating relevant sensorimotor signals needed for consistent task performance [33,34]. Surprisingly, we did not find a difference in dual-task effect between pain status in those with recurrent LBP or between LBP and control groups. This suggests that even though attentional processes were impacted by pain, the cognitive operations that subserve dual-task performance such as procedural processing and goal-selection may be preserved [35]. Pain could be acting like an ongoing, fluctuating task that occupies part of the attentional resources, causing decreases in single task performance and increases in dual-task performance variability, while not affecting the percentage of decrement in task performance in a dual-task condition. Our results, consistent with previous findings, showed that active pain affected task performance even in this young, otherwise healthy cohort. The detriment on arithmetic task performance roughly translates to missing 3 correct subtractions or calculating 3 less responses in one minute than back-healthy individuals. The impact of LBP on daily life and work performance in the long-term may be an issue that warrants further attention.

This was the first study to our knowledge that investigated whether individuals with LBP were able to switch task prioritization when explicitly instructed. We used a dual-task interference paradigm to infer attentional prioritization with and without instructions, and our manipulations were validated by the fact that most individuals performed worse under dual-task conditions compared to single tasks and showed corresponding change in task performance when instructed to switch prioritization between tasks. A few participants demonstrated a facilitation in task performance when dual-tasking, a phenomenon that was occasionally seen in young individuals and may be explained by an overall increase in arousal levels [17]. Inconsistent with our hypothesis, having active LBP or a history of LBP did not impact participants’ ability to switch task prioritization. Previous literature has hinted at a tendency to prioritize the motor task in individuals with LBP [8,11,12]. For example, Etemadi and colleagues reported increased balance recovery velocity during dual-task in persons with LBP [11]. Additionally, Armour-Smith and colleagues reported no change in step length consistency when a cognitive task was introduced to asymptomatic individuals with a history of LBP while the control group was impacted by the additional task [8]. However, if there was prioritization of the motor task, we would expect evidence of a trade-off between tasks with decreased cognitive performance during dual-task conditions in those with LBP compared to controls; this was not the case in these previous studies. The inclusion of explicit task prioritization instructions in our design allows conclusive evidence that young adults with recurrent LBP, regardless of pain status, are able to effectively and voluntarily switch task priorities to a similar degree as their back-healthy counterparts. The question remains: whether persons with LBP prefer to prioritize motor tasks over cognitive tasks. There was no indication of such in our exit interview, with 40% of the LBP-A, 40% of the LBP-R, and 35% of the CTRL participants preferring step width over the arithmetic task. However, two participants with LBP did reveal that “focusing on movement in the step width task helps with the pain”.

Participants with recurrent LBP demonstrated persistent alteration of trunk coordination during narrow-based walking across single and dual-task conditions compared to the back-healthy group that was not influenced by pain status. Young adults with LBP exhibited a more pelvis-dominant and “looser” trunk coordination pattern during narrow-based walking in the frontal plane. There are very few studies on trunk coordination in LBP during dual-task walking, and no previous work could be directly compared with our study, in large part because of discrepancies in methodology and/or study population. Lamoth and colleagues found that patients with chronic LBP had smaller pelvis-thorax relative phase (corresponding to more in-phase, “tighter” control) during treadmill walking across single and dual-task conditions, but the difference was not statistically significant [9]. Armour Smith and colleagues reported no difference in trunk inter-segmental coordination between asymptomatic persons with a history of recurrent LBP and controls during walking turns with and without divided attention [8]. Both studies mentioned above report axial plane coordination, while we chose to focus on frontal plane coordination mostly because of the sensitivity it demonstrates to the demands of the task to walk with a narrow step width [28]. Two previous studies by Rowley and colleagues reported frontal plane movement during a continuous balance-dexterity task. They found decreased trunk coupling (corresponding to less in-phase, “looser” control) in persons with recurrent LBP in symptom remission, which became more similar to the control group under dual-task condition [16,36]. Our findings of a “looser” trunk control were consistent with Rowley’s results in the frontal plane. More studies are needed to disambiguate how trunk coordination is affected by attentional focus, task constraints, and in well-defined subgroups of LBP. Nevertheless, our results provide evidence that the changes in trunk coordination patterns in the recurrent LBP group were independent of pain status and attentional manipulations, potentially indicating habit formation in response to pain. Therefore, the altered trunk coordination appears to have characteristics of a habitual, automatic behavior as it was not impacted by the addition of an attentional-demanding task [37]. This finding also agrees with a previous review suggesting that individuals with LBP adopt a trunk control strategy through a mechanism of reinforcement learning, which persists beyond symptom duration [33].

We chose a within-subject repeated measures design to answer two important questions while reducing variability, thereby increasing the sensitivity: 1) How does active pain perception influence narrow-based walking performance and trunk coordination under dual-task conditions? and 2) does the adapted behavior persist beyond symptom duration in recurrent low back pain? For practical reasons, we always tested participants in pain first, which created the risk of an effect of previous task exposure for the second test administered during remission. While post-hoc analyses did not show evidence of an effect of previous task exposure, our results should be interpreted with caution. Replication studies should aim to randomize the testing sequence in and out of pain. The participants in our study were young, active adults with only mildly disabling recurrent LBP, therefore the findings may not be generalizable to those who are older or have more severe or chronic forms of LBP. In fact, when the effect of aging and chronicity presents itself in those young individuals with recurrent LBP, the alterations in attention will likely become greater.

Two main conclusions can be drawn from the current study. First, the existence of pain negatively impacts cognitive and motor task performance and increases the variability of dual-task performance in young adults with recurrent LBP, compared to that found when the same people are in remission. Second, compared to age-matched back-healthy participants, individuals with recurrent LBP demonstrate a more pelvis-dominant, less in-phase frontal plane trunk coordination pattern during narrow-based walking, regardless of pain status and attentional demands. Additionally, individuals with LBP were able to switch task prioritization when explicitly instructed and did not appear to prioritize the motor task over a cognitive task. These findings provide beginning insights into the relationship between pain status, attention, and motor control in the clinical population of LBP, and have real-world implications in the fields of ergonomics and rehabilitation.

## Data Availability

The data referred to in the manuscript will not be made available online, but may be available upon request to the corresponding author.

## Conflict of interest statement

The authors declare no conflict of interest that could inappropriately influence this work.

## Acknowledgements

This research was supported by the International Society of Biomechanics Matching Dissertation Grant, with matching contribution from the USC Division of Biokinesiology and Physical Therapy. We would also like to thank our participants for their essential contribution to this study.

## Figure Captions

**Supplementary Figure 1.**
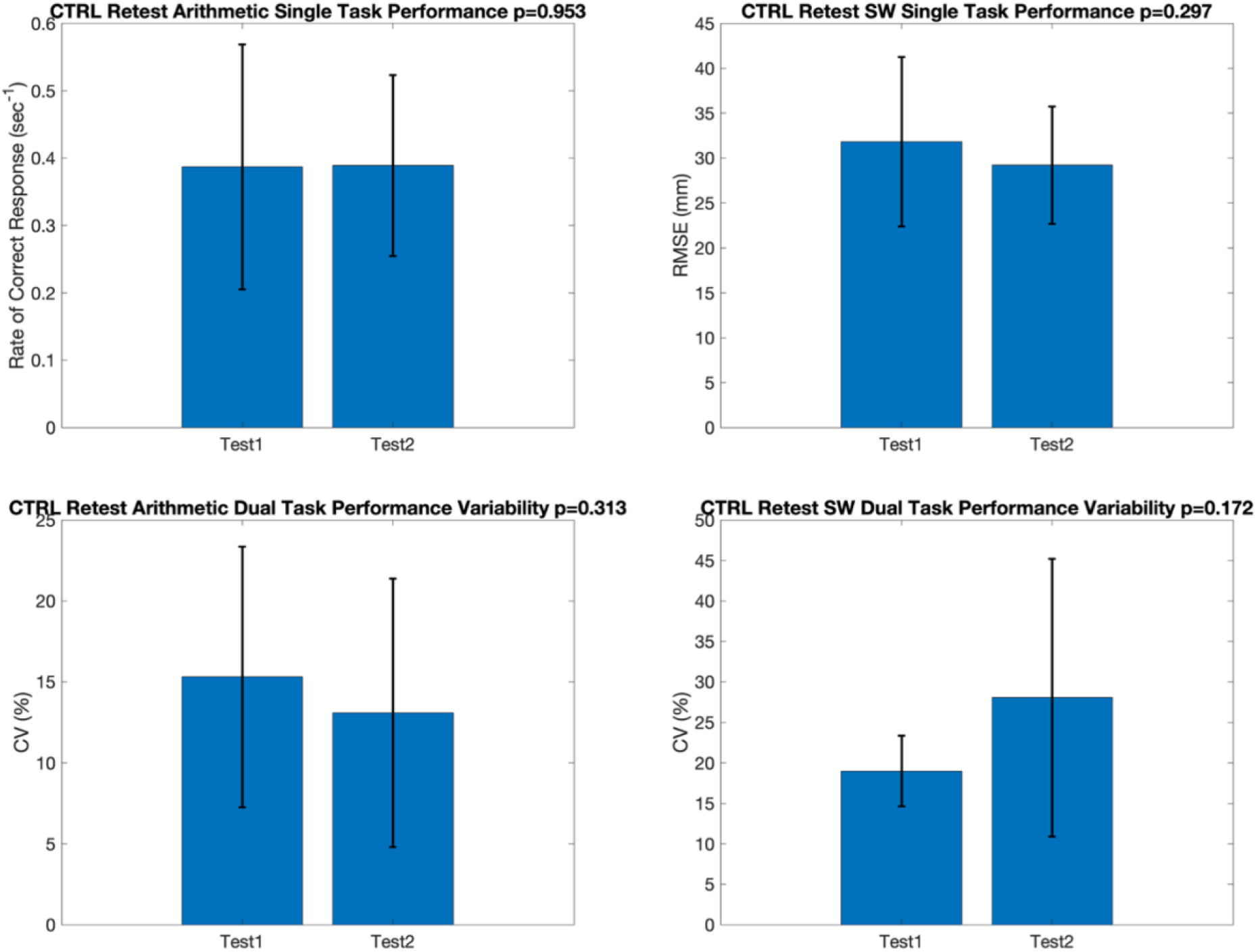
No differences in the comparisons between test-retest of the control group (CTRL) for arithmetic and step width (SW) single task performance and dual-task performance variability, indicating no evidence of practice effect.

**Supplementary Figure 2.**
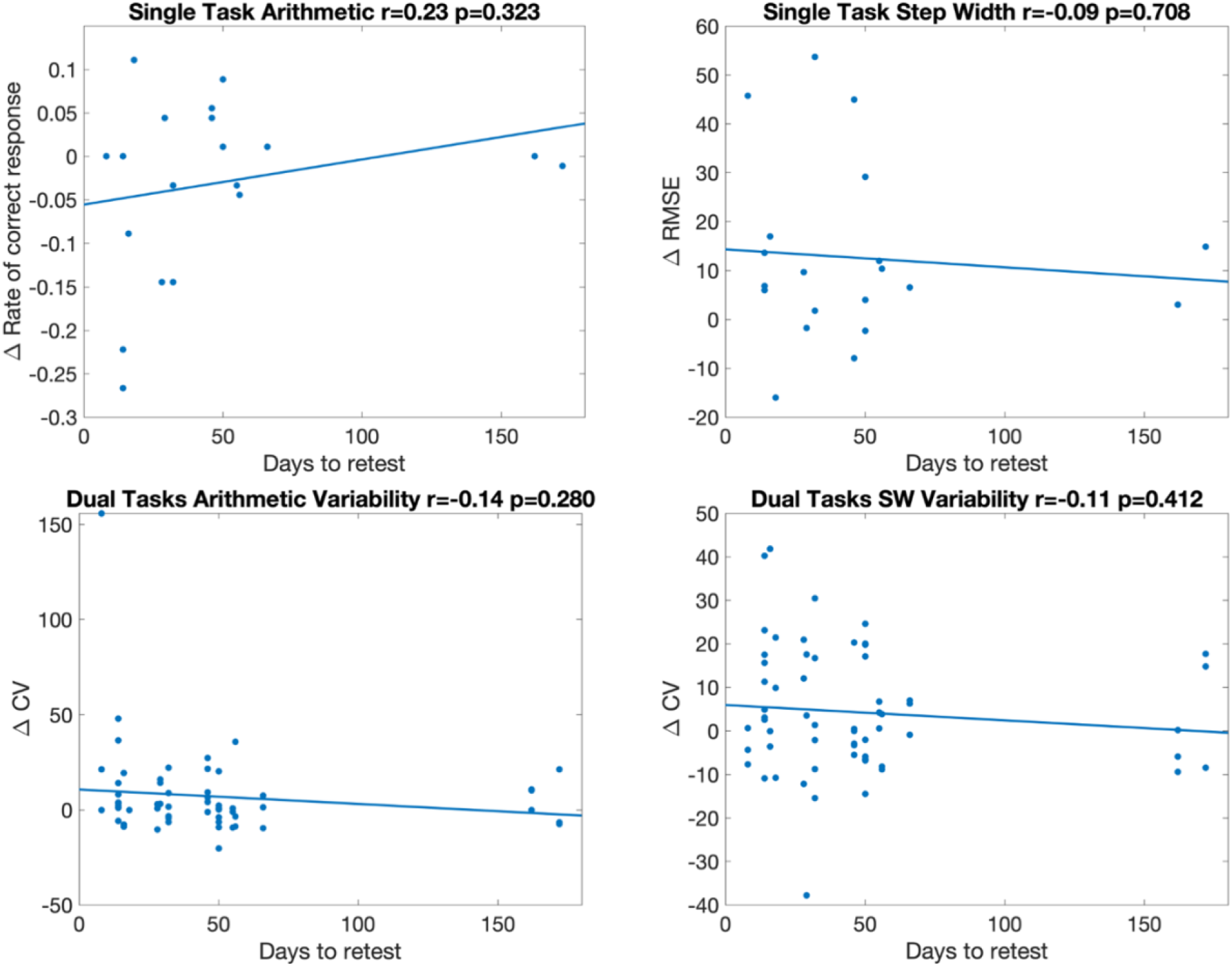
Correlation between days to retest and the changes in arithmetic and step width (SW) single task performance and dual-task performance variability between the in and out of pain testing sessions in individuals with recurrent LBP. There were no relationships for any of those variables, indicating no evidence of improved performance associated with close testing dates.

## Notes

### Competing Interest Statement

The authors have declared no competing interest.

### Clinical Trial

This is a prospective laboratory controlled study without clinical intervention, therefore a clinical trial registration was not needed.

### Author Declarations

University of Southern California Health Sciences Campus institutional review board approved the current study with IRB number: HS-16-00980.

